# Physiotherapists perceived role in managing anxiety in patients with Relapsing-Remitting Multiple Sclerosis: a mixed-methods study

**DOI:** 10.1101/2021.04.06.21254964

**Authors:** Lauren Lucas, Jack Parker

## Abstract

**Objectives:** To explore how physiotherapists manage anxiety in patients with RRMS in community and outpatient settings. To identify further training and research to better develop physiotherapy practice.

**Design:** A mixed-methods design, combining a cross-sectional survey and semi-structured interviews with UK-physiotherapists.

**Participants:** Sixteen UK-registered physiotherapists: 11 in the survey, 5 in the interviews.

**Methods:** To inform the qualitative study, a cross-sectional survey collected data from physiotherapists working in neurology to understand the impact and management of anxiety in people with MS (PwMS) during rehabilitation. Analysis used descriptive statistics and the findings formed the interview guide. Semi-structured interviews with specialist physiotherapists explored barriers and facilitators to managing anxiety, physiotherapy training needs and offered suggestions to develop physiotherapy research and practice. Themes were derived inductively.

**Results:** The survey suggested how PwMS present with anxiety, its impact during rehabilitation, physiotherapy management practices, and physiotherapist skills and training needs. Five semi-structured interviews with specialist physiotherapists expanded on the survey findings and identified five main themes: Understanding the MS journey, modifying assessment and treatment, anxiety management toolbox, lagging behind Musculoskeletal Physiotherapy, and gaining knowledge and skills.

**Conclusion:** Physiotherapists encounter anxiety in PwMS in community and outpatient rehabilitation and perceive they have a role in managing it as it presents. Facilitators included communication, listening skills and opportunities to develop strong therapeutic relationships. Poor training and support, lack of clinical guidelines and limited research evidence were considered barriers. Clinically relevant learning opportunities, interprofessional working, and greater support through clinical supervision is recommended to better develop physiotherapy practice.

**Contribution of the paper:** *Key messages:*

- Physiotherapists perceive they have a role in managing anxiety and psychological wellbeing in PwMS as it presents in community and outpatient neurorehabilitation settings.
- Opportunities to build strong therapeutic relationships through effective communication and listening skills are considered important in being able to manage patient anxiety effectively.
- Poor training and support and a lack of clinical guidelines continue to be barriers in this area.

*New knowledge:*

- Online CPD facilitated through the CSP website and iCSP learning platform may be beneficial to increase competence in this area.
- Further research to understand the MS illness experience in relation to rehabilitation is required to better inform physiotherapy practice and identify appropriate psychologically informed physiotherapy interventions.

## Introduction

MS is a chronic, inflammatory condition affecting the central nervous system and is the most common neurological disorder diagnosed in young adults globally. ^[1,2]^ Around 85% of PwMS will be diagnosed with RRMS at onset, characterised by unpredictable periods of stability (remission) and relapse of symptoms. ^[3]^ As many as 66% of people with RRMS report having anxiety ^[4]^ with greater prevalence in community-based samples. ^[5]^ Anxiety is strongly associated with higher disability levels and significantly impacts pain and fatigue. ^[6]^ It also contributes towards poor treatment adherence, lower social support and social functioning, and poor health behaviours such as smoking and drinking to excess. ^[6]^ In PwMS, anxiety in particular can explain low levels of physical activity, with significant interaction between anxiety and self-efficacy. ^[7]^ Physiotherapists play an essential part in MS management through provision of evidence-based rehabilitation focused on body functions, activities and participation. ^[3]^ Health promotion, to manage risk factors for relapse and disease progression, also constitutes part of the physiotherapist’s role. ^[3]^ However, the impact of anxiety throughout the rehabilitation process is not well known, nor are there clinical guidelines to support physiotherapists in this area.

Few studies have investigated how physiotherapists manage anxiety in their patients, even fewer directly relate to neurological rehabilitation. The recognition of psychosomatic symptoms to differentiate between physical symptoms or those exacerbated by anxiety was considered a key skill in mental health settings. ^[8]^ In vestibular rehabilitation, refined listening and communication skills were perceived to be important to understand the psychosocial complexities of patients, demonstrate empathy, and explain and normalise feelings, in order to build stronger therapeutic relationships. ^[9]^ Approaches used by physiotherapists to manage anxiety are varied, often selected based on clinical experience rather than research evidence. ^[9,10]^ Early research found physiotherapists use core skills of touch and movement; exercise being the most commonly selected treatment, followed by relaxation and breathing retraining. ^[10]^ More recently, greater emphasis has been placed on utilising outcome measures to identify anxiety, with the foundation of treatment being talking and education. ^[9]^ The absence of physiotherapy specific clinical guidelines in the area of anxiety management and psychosocial issues continues to be a significant barrier in clinical practice. ^[9,10]^

In their work, Danielsson et al ^[11]^ and Olund et al ^[12]^ suggest homelikeness as a valuable theoretical framework for the physiotherapy management of anxiety. In this context, the concept of homelikeness relates to the impact of anxiety on the structure of one’s existence, in which the body is a central component. The authors propose that patients experience a disintegration from their body and suggest physiotherapists can address this loss of homelikeness by providing a better understanding and connectedness to the body. In relation to MS where patients experience physical, cognitive and emotional changes contributing to a loss of identity, ^[13]^ this concept may offer a better understanding of the MS illness experience and inform how physiotherapists approach rehabilitation in this patient group.

The aim of this study was to explore how physiotherapists manage anxiety in patients with RRMS in community and outpatient settings and suggest recommendations for future research and practice. Therefore, the research question for this study was:

1. What are the experiences of physiotherapists managing anxiety in patients with RRMS in community and outpatient settings and how can knowledge of these be used to better develop physiotherapy practice?

## Methods

### Design

A mixed-methods study, involving an online survey and semi-structured interviews, was undertaken with UK-based specialist physiotherapists between April and August 2018 as part of an MSc research project. Sequential data collection, with quantitative data collected and analysed to develop the interview schedule, connected data at the analysis stage. As the research problem was conceptualised inductively, through observation in clinical practice and shared experience with other clinicians, greater importance was placed on the qualitative arm of the study. Research ethics approval was granted by the University of Sheffield.

### Sample

190 UK-based Chartered Physiotherapists registered with the Association of Chartered Physiotherapists in Neurology (ACPIN) were invited by email to complete an online questionnaire. Physiotherapists working in neurology at Sheffield Teaching Hospital NHS Trust, Sheffield and Manchester MS Therapy Centres, and a small number of private practices were also invited to participate using gatekeepers in each location. Eligibility criteria included having a registered interest in MS with ACPIN, and currently working or with recent experience of working with PwMS in community and outpatient settings. One reminder email was sent after 3 weeks.

### The survey

A questionnaire was developed using Google Forms, gathering data relating to the impact of anxiety in PwMS during rehabilitation, Physiotherapists skills and techniques to manage anxiety, perceived areas for further physiotherapy training and demographic characteristics. The questionnaire, consisting of predominantly closed questions using ranked-response, multiple choice and Likert scales, was piloted with a sample of 3 physiotherapists representing the target population to enhance validity and reliability. Minor amendments were made to improve question clarity.

### Survey data analysis

Ordinal data was coded using a ‘maximum response score’ based on the total number of responses and analysed in Excel. Results were reported using descriptive statistics. Missing data were excluded from analyses.

### The interview guide

Following quantitative analysis, semi-structured questions were developed to determine how anxiety in PwMS impacts the rehabilitation process, understand the physiotherapists role in managing anxiety, explore training needs and identify areas for further exploration. ^[14]^ The questions, summarised in table 1, were reviewed by a senior academic colleague (JP) and amended to avoid leading or loaded questions. ^[15]^ The guide was also piloted on a physiotherapy colleague impartial to the project, to ensure adequate information was elicited, avoid socially desirable responses and establish an effective interviewing technique. ^[15,16]^

**Table 1.** Summarised interview guide informed by findings from the quantitative survey and literature search

|  |
| --- |
| **Questions informed by the literature search:**<br>- Questions related to the impact of anxiety on the rehabilitation process including perceived barriers<br>- Attitudes towards physiotherapists having a role in managing anxiety and psychological wellbeing in PwMS during the rehabilitation process<br>- How the profession can move to consider psychological aspects of health and wellbeing in PwMS and other neurological conditions/in neurological rehabilitation settings |
| **Questions informed by the quantitative survey:**<br>- Exploring prioritisation of anxiety, depression and stress.<br>- Having high expectations of others vs. what patients expect from physiotherapy in relation to physical and psychological wellbeing<br>- Issues associated with trouble concentrating, worry and avoidance during the rehabilitation process and how these are managed clinically<br>- Exploration of physiotherapists feeling comfortable asking about mental health but less comfortable acting on disclosures<br>- Effective treatment approaches to managing anxiety in PwMS and useful interventions/training/skills for improving in this area in the future<br>- Exploration of psychological content of training and moving forward in this area |

### The interviews

Purposeful sampling of Physiotherapists involved in the quantitative study were invited via gatekeepers to participate in the interviews. Eligibility included physiotherapists working in neurology with current or previous experience in community or outpatient neurorehabilitation. Physiotherapists working in inpatient neurorehabilitation were excluded. Face-to-face and telephone interviews were offered to enhance participation in the study. Informed consent was gained in written format prior to the interview. Interviews lasted between 22 and 49 minutes in total. Collection of demographic information provided additional context to the data. Interviews were transcribed verbatim and anonymised by the interviewer.

### Data analysis

Data were analysed using Braun and Clarkes thematic analysis. ^[17]^ Complete coding, using NVivo 12 software, identified all incidences deemed interesting or relevant to the research question. ^[15]^ With limited existing research on the topic, thematic analysis at a latent level examined the underlying assumptions, ideas and ideologies shaping the semantic substance of the data. ^[17]^ This involved producing researcher-derived codes to identify implicit, rather than explicit meaning. ^[15]^ An inductive approach to analysis ensured themes were grounded in participants responses. ^[18]^ Themes and supporting quotes are detailed in Table 2. Qualitative data were reported in line with the Standards for Reporting Qualitative Research (SRQR). [19]

**Table 2.** Themes, subthemes and examples of quotes from qualitative interviews.

|  |
| --- |
| **CURRENT PRACTICE** |
| **Theme 1: Understanding the MS Journey** |
| **Subtheme: Physiotherapist Role** |
| *"It's being able to know when it's not actually for you and you need to hand it over" (PT1)* |
| *"I guess for neuro there's that really good understanding of the acceptance journey that people go on. Because that's when we see anxiety come out, that's when we see depression come out, that's when we see anger and frustration and those, and probably that difference between what's "normal" anxiety and depression and moving on that journey of grief and acceptance and what's dysfunctional and potentially dangerous" (PT5)* |
| **Subtheme: Patient's expectations** |
| *"I think MS patients' expectation of physio in general is quite high because they see physio as the thing that's going to get them better because there isn't a wonder drug. With MS it isn't like that, they, they hang everything on their physio." (PT2)* |
| *"We have some people who come expecting somebody to do something to them...and convincing people to continue exercises, or other interventions anyway, when they're not here can be quite difficult" (PT3)* |
| **Theme 2: Modifying assessment and treatment** |
| **Subtheme: Impact of anxiety** |
| *"I think the anxiety, if people have that when they come in for whatever reason, that can really sort of inhibit how much they take on." (PT3)* |
| *"How good are we at you know, having our own strategies to work with that population, remain positive and not get to that point of burn out or, stress or, beating yourself up because you think you haven't delivered a good session" (PT5)* |
| **Subtheme: Management strategies** |
| *"I'm not sure that the GP would always be the best person to help deal with [anxiety], but they might have access to local services to help that patient...the MS nurses are really good sources of information and support for patients." (PT3)* |
| *"I think that signposting to the more psychological therapies, I think we have to be really knowledgeable about those things." (PT5)* |
| **Theme 3: Anxiety management toolbox** |
| **Subtheme: Skills and attributes** |
| *"I think you can alleviate somebody's worry just by talking to them." (PT2)* |
| *"I think the ability to empathise and to listen and to give a degree of understanding and gain a rapport, its all, it's all involved in being able to effectively communicate both verbally and non-verbally I guess" (PT4)* |
| **Subtheme: Barriers and facilitators** |
| *"its lack of confidence and feeling like they haven't got the tools in their tool kit to be able to give the appropriate advice to people." (PT2)* |
| *"I think if you were going into it and you were newly qualified, I think you wouldn't want to be delving into that area because you probably don't have that experience to go off" (PT4)* |
| *"Perhaps there's something about us looking in, I talked about supervision. If you look at psychology, social work or OT\*, they're really good at addressing their own wellbeing within supervision." (PT5)* |
| **MOVING FORWARDS** |
| **Theme 4: Lagging behind MSK\*** |
| *"They're getting better at that in MSK than we possibly are in neuro...Pete O'Sullivan people like that are doing courses where you're listening to the patient's story and what that is actually telling you about how that's impacting them, and I think that's really important" (PT2)* |
| *"If you look within chronic pain there is evidence that psycho-education changes people's physical symptoms or experience of pain. If we can do that in chronic pain, we can do that in every condition." (PT5)* |
| **Theme 5: Gaining knowledge and skills** |
| **Subtheme: Interprofessional working** |
| *"I'm sure that the MS nurses will do absolutely loads of psychological support, because you know in their role, so actually there's probably a lot that we could learn and a lot that we could share between us." (PT1)* |
| *"Learning from psychology, I've learned heaps from working with psychologists" (PT5)* |
| **Subtheme: Physiotherapy training** |
| *"At an undergrad level it needs much more emphasis, about we need to look at the whole person and looking at the evidence now that supports how the psychological impact of things affects physical symptoms, we're no longer just treating physical symptoms were treating the whole patient." (PT2)* |
| *"more of a guidance package than a training package sort of you know...coming sort of centrally as part of CSP\*, guidance or case studies for example on ICSP" (PT3)* |
| **\*CSP = Chartered Society of Physiotherapy** |
| **\*MSK = Musculoskeletal** |
| **\*OT = Occupational Therapy** |

## Results

### Quantitative phase

Eleven respondents, characterised in Table 3, completed the survey with one question missing a response. All participants were female and clinical experience ranged between 1 − 38 years. Of the respondents, 82% worked in community, outpatient or private practice neurology settings and 18% worked in other areas but had experience in neurology. 45% (n = 5) had postgraduate training in psychological interventions. Of those, 60% had training in motivational interviewing (MI), 20% in cognitive behavioural therapy (CBT), and 20% had informal training.

**Table 3.** Demographic characteristics of survey respondents

|  |  | Frequency<br>(n = 11) |
| --- | --- | --- |
| **Gender** | Female | 11 |
|  | Male | 0 |
| **Type of Work** | Full time | 4 |
|  | Part time | 7 |
| **Area of work** | Outpatient clinic | 5 |
|  | Community | 1 |
|  | Private practice | 3 |
|  | Other (with Neuro experience) | 2 |
| **Region of work** | South West | 3 |
|  | East of England | 1 |
|  | London | 2 |
|  | Yorkshire and the Humber | 2 |
|  | Not reported | 3 |
| **Years of experience** | 1 – 10 years | 1 |
|  | 11 – 20 years | 5 |
|  | 21 – 30 years | 2 |
|  | 30+ years | 3 |
| **Range of experience (years)** | 1 – 38 |  |
| **Mean years of experience** | 22.9 |  |
| **Median years of experience** | 20 |  |
| **Post graduate training in psychology** | Yes | 5 |
|  | No | 6 |
| **Type of training** | CBT | 1 |
|  | Motivational Interviewing | 3 |
|  | Informal training | 1 |

#### How people with RRMS present

Respondents reported that stress (63%), depression (59%) and sadness (57%) were most frequently encountered in PwMS however, anxiety and depression were prioritised as most important to manage during physiotherapy interventions (Figure 1). Trouble concentrating (56%) and worry (53%) were common emotional symptoms observed, followed by avoidance (41%). To understand the impact of anxiety in relation to rehabilitation, physiotherapists were asked to consider negative patterns associated with anxiety and the extent to which they arose in PwMS clinically. High expectations of others (80%), dwelling on the past (70%), over generalising (70%) and labelling (70%) were most frequently observed.

**Figure 1.**
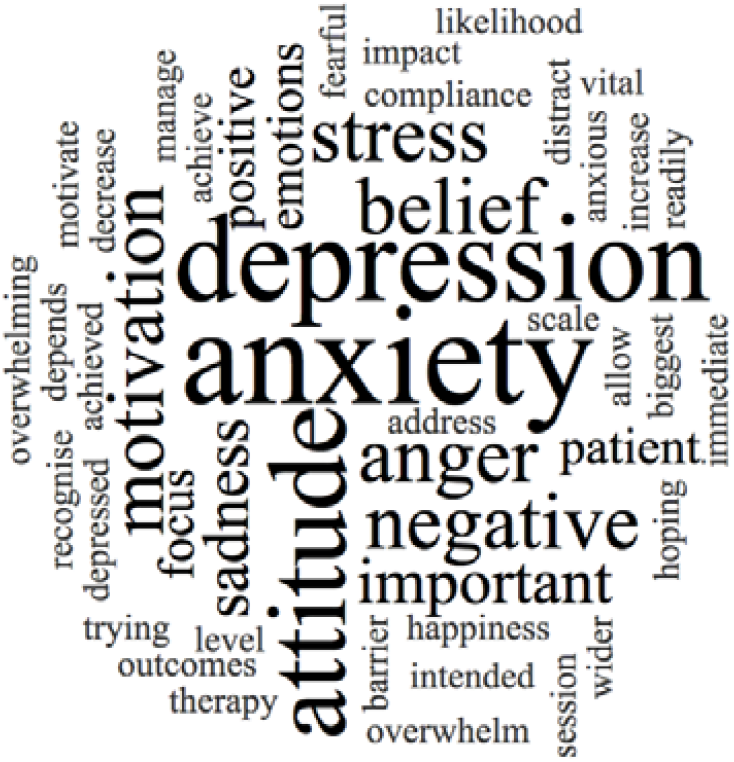
Factors most important to manage during physiotherapy treatments based on free text answers.

#### Managing anxiety in clinical practice

Respondents identified anxiety in PwMS through informal conversations with other members of the team (91%), however MDT meetings were not selected by any participants. Self-reporting from patients constituted 91% of the responses and most participants (82%) felt clinical experience was a significant factor for identifying anxiety. All participants felt confident asking about mental health problems, however responses were widely spread in relation to acting on, or dealing with a disclosure (Figure 2). Table 4 details treatment approaches for anxiety used by the respondents. Goal setting (91%) was most common, followed by relaxation (82%) and reflective listening (73%). Despite only 3 participants receiving training in MI, it was a treatment approach selected by 7 participants (64%).

**Figure 2.**
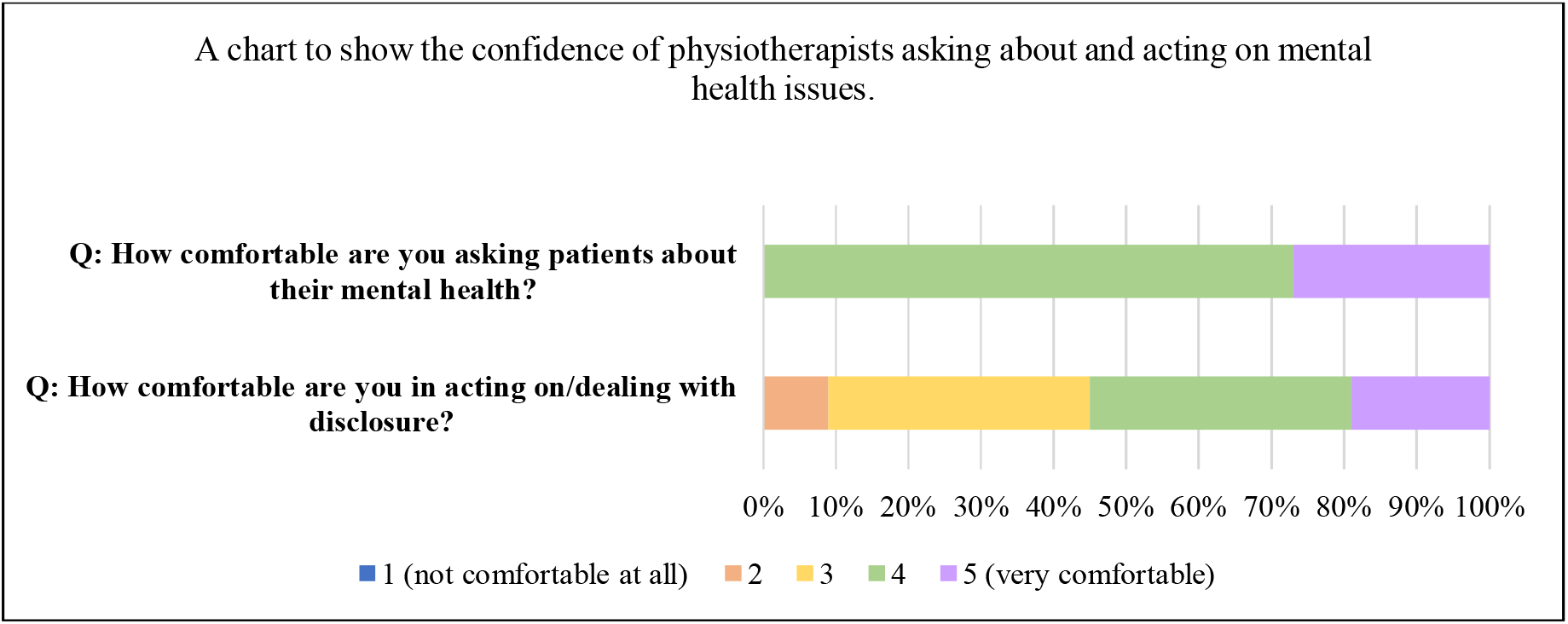
Percentage of survey responses for questions around confidence asking about mental health and acting on disclosures (n=11)

**Table 4.** Table summarising treatment techniques used to manage anxiety.

| Treatment technique | Frequency (n=11)<br>% (n) |
| --- | --- |
| **Goal setting** | 91% (10) |
| **Relaxation** | 82% (9) |
| **Reflective listening** | 73% (8) |
| **Motivational Interviewing** | 64% (7) |
| **Workbooks / paper resources** | 64% (7) |
| **Coping Strategies** | 55% (6) |
| **Mindfulness** | 55% (6) |
| **Positive reinforcement** | 45% (5) |
| **Imagery / visualisation** | 36% (4) |
| **Self-talk** | 27% (3) |
| **Cognitive Behavioural Therapy (CBT)** | 18% (2) |
| **Acceptance and Commitment Therapy (ACT)** | 0% (0) |

#### Skills and training required to manage anxiety in patients

Effective communication (89%) and listening skills (88%) were deemed most important to managing anxiety. However clinical experience (61%) and working towards goals (59%) were less important, despite goal setting being the most frequently used treatment technique for anxiety. Involving the MDT (55%) and offering advice (39%) was least important. Figure 3 details participants responses to identifying interventions useful for managing anxiety and indicating those where additional training would be useful. Interestingly, MI scored similarly in its value as an intervention in clinical practice (57%) and additional training (58%). Similar incidence was found for mindfulness, scoring 53% and 57% respectively. Most participants (82%) agreed psychology was an important part of physiotherapy training. However, responses were skewed regarding effectiveness of undergraduate training in providing skills to successfully navigate patient psychological wellbeing.

**Figure 3.**
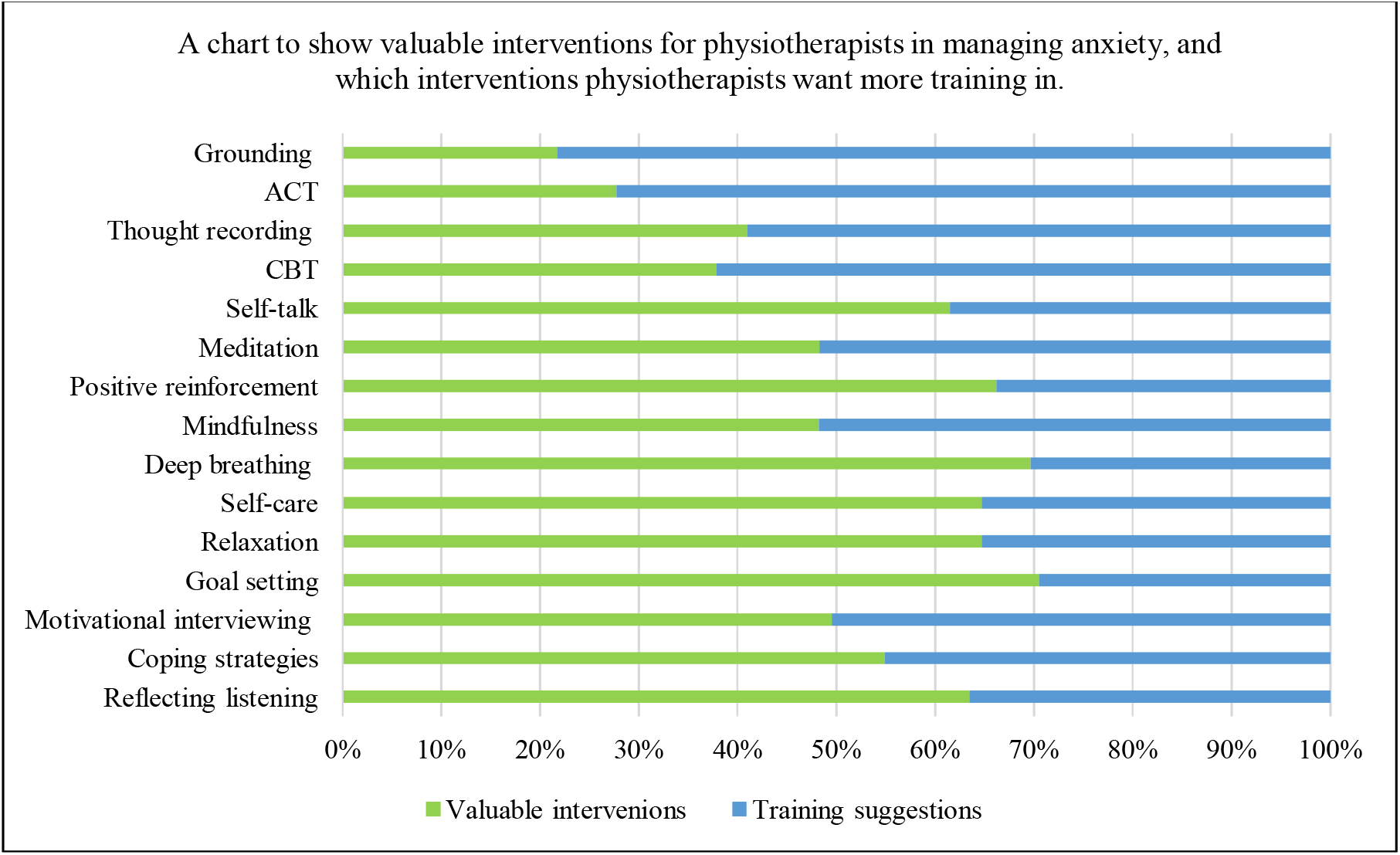
A chart summarising the priority in which interventions are considered valuable to physiotherapists to manage anxiety, compared to which interventions are prioritised for additional training.

### Qualitative phase

Five physiotherapists, characterised in Table 5, participated in semi-structured interviews. All participants worked in neurology with a greater proportion having experience in MS therapy centres (n = 3). Three participants worked in community and outpatient settings whilst two worked in clinic settings. Three participants received informal psychological training. Figure 4 displays a visual representation of the overarching themes, themes and subthemes. A single direction arrow establishes hierarchical relationships; bi-directional arrows indicate close lateral relationships; a dashed line signals a tentative relationship.

**Table 5.** Demographic characteristics of interview participants

|  |  | **Frequency**<br>**(n = 5)** |
| --- | --- | --- |
| **Gender** | Female | 5 |
|  | Male | 0 |
| **Work setting** | NHS | 1 |
|  | Private practice | 4 |
| **Experience working in MS therapy centre (current/past)** | Yes | 3 |
|  | No | 2 |
| **Hours of Work** | Full time | 4 |
|  | Part time | 1 |
| **Area of work** | Outpatient clinic only | 2 |
|  | Community & Outpatient clinic | 3 |
| **Frequency seeing PwMS** | Daily | 2 |
|  | Weekly | 2 |
|  | Fortnightly | 1 |
| **Post graduate training in psychology** | Yes | 3 |
|  | No | 2 |
| **Type of training** | Informal (in-service training) | 3 |

**Figure 4.**
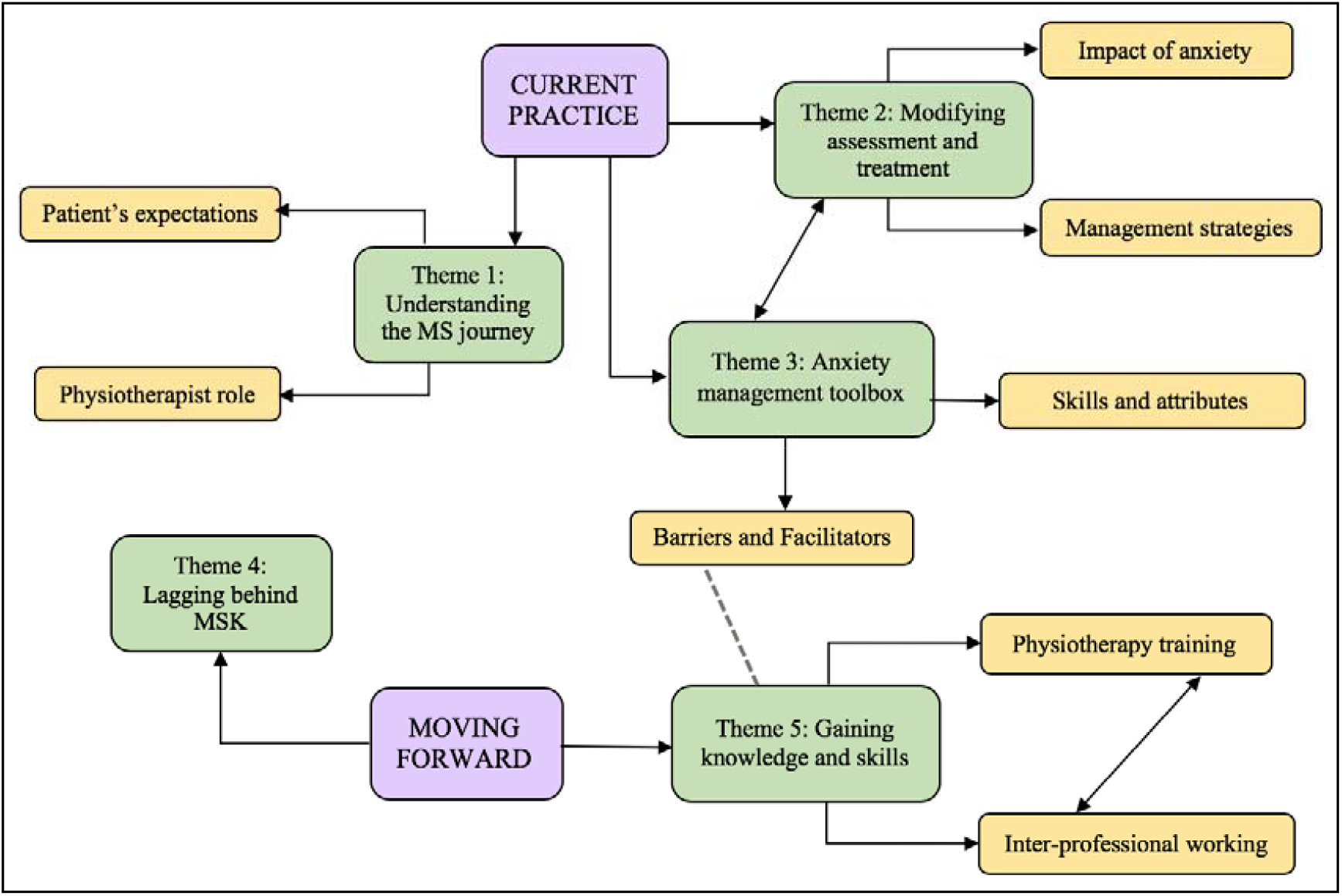
Visual map of participant overarching themes, themes and subthemes

#### Theme1: Understanding the MS Journey

A common idea was understanding the MS journey significantly differs to other neurological conditions. Within this theme participants discussed their role when treating PwMS, patient expectations and the challenges they face.

##### Subtheme: Physiotherapist Role

In addition to facilitating movement and function, physiotherapists are required to support acceptance and adjustment following a life-altering health event or diagnosis. This requires understanding the MS illness experience, the impact on social, family and life roles and manage anxieties related to these factors. Despite being well placed to take an active role in managing anxiety in clinical interactions, they recognised the importance of working within their scope of practice and seeking support elsewhere in more challenging situations.

##### Subtheme: Patient’s Expectations

A significant challenge was the importance placed on physiotherapy, from patients and other professionals, as a way to manage symptoms in the absence of any curative treatment. Misconceptions around physiotherapy necessitating hands-on treatment challenged productive self-management discussions. As the measure of success is inherently different in progressive neurological conditions, physiotherapists often felt like they were not helping their patients therefore not meeting patient expectations.

#### Theme 2: Modifying assessment and treatment

Participants acknowledged the many layers of anxiety that PwMS can face and recognised it exists within physiotherapy treatment sessions. In identifying the impact of these anxieties, participants reflected on how they manage it as it presents, reflected in the following subthemes.

##### Subtheme: Impact of Anxiety

All participants reflected anxiety blocked treatment progression, particularly through greater time discussing anxiety related issues and the implications of this on physical progression. Participants felt physiotherapy itself contributed towards anxiety, referencing challenges to physical ability, indicating the importance of exploring the source and reflexively modifying treatment. Anxiety was detrimental to self-efficacy, affecting concentration and retention of information, therefore impeding self-management. An interesting insight was the impact of anxiety on physiotherapists, with clinicians feeling less competent in their clinical skills and noting feelings of inefficacy.

##### Subtheme: Management Strategies

All participants used treatment planning as an anxiety management strategy, ranging from task specific training and goal setting, to changing the layout of the room. Education strategies varied between education relating to the physiotherapy process and psychoeducation to normalise anxiety symptoms. There were mixed views around using psychological interventions as part of physiotherapy treatment. Signposting and onward referrals were emphasised, with MS nurses being a more valuable resource than GP’s. All participants adopted anxiety management strategies within their clinical practice, however expressed difficulty meeting the psychological needs of their patients.

#### Theme 3: Anxiety management toolbox

Theme two and three are closely related as the process of managing anxiety does not solely rely on interventions and clinical approaches, but the skills required to employ them. The subthemes ‘Skills and attributes’ and ‘barriers and facilitators’ build upon what is required for physiotherapists to create an anxiety management toolbox.

##### Subtheme: Skills and attributes

Interpersonal skills such as communication skills, active listening, and empathy to form stronger therapeutic relationships, underpinned the anxiety management toolbox. Strong therapeutic relationships allow patients to open up about their worries allowing physiotherapists to alleviate concern, signpost accordingly and set specific rehabilitation goals. Clinical reasoning was important to evaluate the success of interventions, providing opportunity to question why progress may not as expected and address underlying issues, such as anxiety, posing as barriers.

##### Subtheme: Barriers and Facilitators

All participants felt strongly that undergraduate training did little to prepare new graduates for the challenges they are likely to face managing psychological wellbeing. Barriers included a lack of confidence and skills, inadequate undergraduate and postgraduate training opportunities and clinical supervision. Facilitators included clinical setting and team support, having access to psychology input, and opportunity to build a therapeutic relationship.

#### Theme 4: Lagging behind MSK

Participants felt neurological physiotherapy may be behind, in both research and practice, when looking at the link between physical and mental health. Particular reference was made to evidence supporting the psychosocial management of chronic pain within physiotherapy and how these principles could be applied to the neuro setting. Using patient narrative over gold standard research methods to understand the patient story was also deemed important in the context of neuro-research.

#### Theme 5: Gaining Knowledge and skills

Suggestions to gain knowledge and skills to effectively manage anxiety in PwMS were detailed in the following subthemes.

##### Subtheme: Interprofessional Working

Interprofessional working was strongly woven through all of the interviews. This included working with psychology, MS nurses and occupational therapists, and the recommendation of a network for therapists interested in MS. Despite recognising that interprofessional working was important, it was noted that this opportunity may not be available to all therapists.

##### Subtheme: Physiotherapy Training

Several suggestions to support physiotherapy training at undergraduate and postgraduate level were made. These included training in the use of psychologically informed physiotherapy and opportunities to enhance communication and listening, joint training with psychology and MS nurses, and utilising interactive, online training platforms through the CSP.

## Discussion

This study explores physiotherapists’ perspectives on the impact and management of anxiety in PwMS. Participants felt it was part of their role to manage psychological and emotional wellbeing throughout the rehabilitation process and described the impact of anxiety on physiotherapy interventions. In the absence of current evidence or treatment guidelines, physiotherapists feel underprepared and often lack the confidence to do this effectively in clinical settings.

Anxiety was found to affect self-efficacy, participation, rehabilitation progression and treatment adherence, with many factors influencing the presence of anxiety in PwMS; factors directly relating to the disease, psychosocial issues or the rehabilitation process itself. With the introduction of homelikeness theory to explain the presence of anxiety in the body, ^[11,12]^ qualitative interviews highlighted both physical and social influences contributing towards feeling unhomelike. In the context of chronic illness, Svenaeus ^[20]^ suggests illness brings about feelings of unhomelikeness in the social world, reflected in the findings of this study. Similarly, Jingrot and Rosberg ^[21]^ discuss fatigues impact on homelikeness, fatigue being a common experience for PwMS and directly impacting upon anxiety. Despite this term not arising during the study, physiotherapists believed it was part of their role to alleviate the presence of anxiety, therefore mitigating unhomelikeness, using skills they felt were within their scope of practice.

Verbal and non-verbal communication, empathy and listening, were identified by Physiotherapists as key to managing anxiety in PwMS through developing strong therapeutic relationships. Similarly to vestibular rehabilitation, ^[9]^ participants emphasised development of listening and communication skills to build trusting relationships, allowing patients to open up about their problems. A recent study identified the intimate nature of the patient/physiotherapist relationship; patients disclosing personal information related to their wellbeing outside of the therapy setting, is one that is important for engagement in the therapeutic relationship. ^[22]^ However, there were a number of limitations to this, including its application to patients with neurological conditions due to therapeutic relationships extending to family members. Interestingly in this study, managing the wider impact of anxiety within family contexts was not discussed. Therefore, the wider extent to which the therapeutic relationship is utilised in community and outpatient neurological rehabilitation is unknown and may benefit from future research.

Despite feeling comfortable asking about mental health, findings indicated that physiotherapists were less comfortable acting on what might be disclosed, largely due to a perceived lack of skills to navigate difficult conversations. This is reflected in musculoskeletal physiotherapy where physiotherapists report inadequate skills, time, support and education to effectively assess and manage the complex psychosocial factors associated with chronic pain. ^[23,24]^ Being long-term conditions, the physiotherapy management of chronic pain and MS is not dissimilar, with guidelines promoting self-management. ^[25-27]^ However, the former places greater emphasis on concurrently managing both mental and physical wellbeing, with a move towards physiotherapists providing psychologically informed treatments such as CBT and ACT. This has been found acceptable to patients ^[28,29]^ and physiotherapists delivering these treatments, ^[30,31]^ bringing into question why this expansion of the physiotherapist’ role has not been considered in neurological rehabilitation. Few participants in this study used psychologically informed interventions in their practice. Although quantitative findings indicated that training in ACT and CBT may be beneficial, qualitative findings suggested that physiotherapists need only to expand their knowledge in these areas to support patient decision making. Due to the small sample size, further research would be needed to gain a broader understanding of how beneficial psychologically informed physiotherapy would be in community and outpatient neurorehabilitation.

In addition to a lack of skills, a significant barrier to physiotherapists feeling competent was inadequate training, at both undergraduate and post-graduate level. This reflects findings from other clinical settings where physiotherapists felt their training did not equip them with skills to meet the psychological needs of their patients. ^[32,33]^ Qualitative interviews suggested several recommendations for improvements in this area, for example the use of narrative to develop case studies and patient stories such as those used in chronic pain research. This study highlighted physiotherapists understand the impact of uncertainty and loss of identity associated with chronic disease, however, lack the skills to confidently manage the illness experience during rehabilitation. Narrative has been used to understand the lived experience of pain-related fear to better inform its physiotherapy management, reconceptualising assessment and treatment approaches as a result. ^[34,35]^ A similar approach may be beneficial within neurological physiotherapy to better understand the multifaceted biopsychosocial aspects of MS that directly impact rehabilitation.

Interprofessional working and training was another recommendation to move forward in this area. However, opportunities for interprofessional working appear to be limited in community and outpatient settings, a finding reflected in the vestibular rehabilitation setting. ^[9]^ In psychiatry and pharmacology, the use of virtual learning environments has been shown to be effective at facilitating interprofessional learning at a post-graduate level. ^[36,37]^ This study indicated that physiotherapists may be open to online CPD facilitated through the CSP website and iCPS learning platform. Therefore, this approach may support interprofessional learning with psychologists and MS specialist nurses, using clinically relevant scenarios, case studies and patient stories. In addition to post-graduate training, ongoing supervision and reflective practice consolidates skills to manage psychological wellbeing effectively. Poor support and ineffective supervision can contribute towards burnout in physiotherapists, a concept directly affected by feelings of inefficacy. ^[38]^ The themes ‘impact of anxiety’ and ‘patient expectations’ introduced the idea of inefficacy, where physiotherapists reported the challenges of working with MS and self-criticism of their clinical skills. Interprofessional supervision may support greater skill acquisition and offer alternative perspectives to solving clinical problems.

This study had some limitations. Poor survey response significantly limits the generalisability of the findings. Moreover, findings from both phases may be biased towards gender, years of clinical experience and setting as all participants were female averaging 22 years clinical experience. A greater proportion also had experience working in MS Therapy Centres thus developing specialist skills. The study findings cannot accurately reflect the experience of male physiotherapists or lower grade rotational physiotherapists with less clinical experience in neurology.

## Conclusion

This study demonstrates physiotherapists perceive their role to include managing anxiety in PwMS, with anxiety impacting self-efficacy, participation and engagement with self-management strategies in community and outpatient rehabilitation. A lack of confidence, skills, opportunity to obtain skills, clinical guidelines and clinical support remain ongoing barriers to developing in this area. There is limited research investigating how physiotherapists manage psychological wellbeing in patients with long-term neurological conditions. Moreover, little is known about the skills required to manage the wider impact of neurological conditions, for example family and carer wellbeing. Further research is required, using methodologies that accurately reflect the patient journey to inform better understanding of the MS illness experience directly related to rehabilitation. This may lead to the development of psychologically informed interventions relevant to neurological physiotherapy and inform appropriate guidelines to support physiotherapists to manage anxiety across different clinical settings.

## Data Availability

The authors confirm that the data supporting the findings of this study are available within the article.

## Ethical approval

The University of Sheffield School of Health and Related Research (ScHARR) Research Ethics Committee approved this study. All participants gave written informed consent before data collection began.

